# Persistent while declined neutralizing antibody responses in the convalescents of COVID-19 across clinical spectrum during the 16 months follow up

**DOI:** 10.1101/2021.09.18.21263550

**Authors:** Yang Yang, Minghui Yang, Yun Peng, Yanhua Liang, Jinli Wei, Li Xing, Liping Guo, Xiaohe Li, Jie Li, Jun Wang, Mianhuan Li, Zhixiang Xu, Mingxia Zhang, Fuxiang Wang, Yi Shi, Jing Yuan, Yingxia Liu

## Abstract

Elucidation the kinetics of neutralizing antibody response in the coronavirus disease 2019 (COVID-19) convalescents is crucial for the future control of the COVID-19 pandemic and vaccination strategies. Here we tested 411 sequential plasma samples collected up to 480 days post symptoms onset (d.a.o) from 214 convalescents of COVID-19 across clinical spectrum without re-exposure history after recovery and vaccination of SARS-CoV-2, using authentic SARS-CoV-2 microneutralization (MN) assays. COVID-19 convalescents free of re-exposure and vaccination could maintain relatively stable anti-RBD IgG and MN titers during 400∼480 d.a.o after the peak at around 120 d.a.o and the subsequent decrease. Undetectable neutralizing activity started to occur in mild and asymptomatic infections during 330 to 480 d.a.o with an overall rate of 14.29% and up to 50% for the asymptomatic infections. Significant decline in MN titers was found in 91.67% COVID-19 convalescents with ≥ 50% decrease in MN titers when comparing the available peak and current MN titers (≥ 300 d.a.o). Antibody-dependent immunity could also provide protection against most of circulating variants after one year, while significantly decreased neutralizing activities against the Beta, Delta and Lambda variants were found in most of individuals. In summary, our results indicated that neutralizing antibody responses could last at least 480 days in most COVID-19 convalescents despite of the obvious decline of neutralizing activity, while the up to 50% undetectable neutralizing activity in the asymptomatic infections is of great concern.

## Introduction

Severe acute respiratory syndrome coronavirus 2 (SARS-CoV-2) was the seventh member of coronaviruses that has been found to infect human beings in late December, 2019, in Wuhan, China^1,2^. The pandemic of SARS-CoV-2 is ongoing and has caused over 209 million human infections with over 4.4 million fatal cases worldwide until now (https://covid19.who.int/). Notably, asymptomatic infections account for up to 40% of all the SARS-CoV-2 infections, and silent transmission during the pre-symptomatic and asymptomatic stages are responsible for more than 50% of the overall attack rate in COVID-19 outbreaks^3-6^. Therefore, antibody-mediated protective immunity induced either by natural infection or vaccination is of great importance for the control of COVID-19 pandemic.

Neutralizing antibodies (NAbs) rapidly rise after infection and maintained for years to decades by long-lived plasma and memory B cells in most acute viral infections, playing vital roles in the viral clearance and protection against viral diseases^7^. Studies have shown that the antibody response induced by coronavirus in human beings tends to wane over time, and varies among types of coronaviruses and the severity of the disease^8^. Seasonal human coronaviruses including HCoV-229E, HCoVOC43, HCoV-NL63, and HCoV-HKU1 induce short-lasting antibody response, and reinfection with the same seasonal coronavirus occurred frequently at 12 months after infection^9^. However, the two coronavirus that cause severe disease in human beings including SARS-CoV and Middle East Respiratory Syndrome coronavirus (MERS-CoV) induce stronger and more durable antibody responses up to 3 years^8,10-14^. Recent data from animal models, therapeutic use of neutralizing monoclonal antibodies and convalescent plasma suggest the critical role of neutralizing antibody in controlling SARS-CoV-2 infection and reinfection^15-22^, while a key question yet to be addressed is whether COVID-19 convalescents could maintain long-lasting antibody mediated protective immunity. Studies have set out to investigate the dynamic change and longevity of both binding and neutralizing antibody response in COVID-19 convalescents^14,23-31^, and the results in a very recent study indicated that neutralizing antibody response in COVID-19 convalescents could maintain up to 300 days after symptoms onset (d.a.o) accompanied by waning of neutralizing antibody titers^29-31^.

Limitations of existing studies included either limited number of cohort, or short duration of follow up, enzyme-linked immunosorbent assay (ELISA) or pseudoparticle neutralization based detection of neutralizing antibody response. Moreover, possible re-exposure to SARS-CoV-2 in the pandemic area of COVID-19 could significantly influence the dynamics and longevity of NAb titers in COVID-19 convalescents. Owing to the highly efficient control of the COVID-19 endemic in China, cohort in China offers ideal model to evaluate the natural dynamics and longevity of neutralizing antibody responses after SARS-CoV-2 infection. In this study, we investigated the dynamics and longevity of NAb titers up to 480 d.a.o from 214 convalescents of COVID-19 across clinical spectrum without any further exposure history after recovery and vaccination of SARS-CoV-2, using authentic virus based microneutralization assay, and tested the neutralizing activity against the circulating variants utilizing SARS-CoV-2 pseudotype neutralization assays.

## Materials and Methods

### Patient information and sample collection

Subjects presented in this study were laboratory confirmed COVID-19 patients using quantitative real-time PCR (Mabsky Biotech Co.) and admitted to our hospital during January 11 to April 18, 2020 (N=214). These individuals were further divided into severe (including critically ill and severe cases), mild (including mild and moderate patients) and asymptomatic group based on disease severity categorized according to the China National Health Commission Guidelines for Diagnosis and Treatment of SARS-CoV-2 infection. Clinical information and laboratory result were collected at the earliest time-point after hospital admission. Blood samples were collected from the enrolled patients during hospitalization and follow-up. Individuals with any further exposure history after discharge or vaccination of SARS-CoV-2 were ruled out during follow-up. The study protocol was approved by the Ethics Committees of Shenzhen Third People’s Hospital (2020-010). Written informed consents were obtained from all patients. The study was conducted in accordance with the International Conference on Harmonisation Guidelines for Good Clinical Practice and the Declaration of Helsinki and institutional ethics guidelines.

### Disease severity classification

Disease severity classification was evaluated according to China National Health Commission Guidelines for Diagnosis and Treatment of SARS-CoV-2 infection (seventh version) as previously reported^32,33^. In brief, laboratory confirmed patients with fever, respiratory manifestations and radiological findings indicative of pneumonia were considered moderate cases. Laboratory confirmed patients who met any of the following were considered to have severe COVID-19: 1) respiratory distress (respiration rate ≥ 30/min; 2) resting oxygen saturation ≤ 93%, or 3) arterial oxygen partial pressure (PaO_2_) / fraction of inspired oxygen (FiO_2_) ≤ 300 mmHg (1 mmHg = 0.133 kPa). Laboratory confirmed patients who had any of the following were considered critically ill: 1) respiratory failure requiring mechanical ventilation, 2) shock, or 3) failure of other organs requiring intensive care unit (ICU). The asymptomatic cases were defined as previously reported^34^.

### Cell lines and viruses

African green monkey kidney Vero cell (ATCC, CCL-81) were obtained from ATCC, and 293 cells stably expressing ACE-2 (293-ACE-2) were obtained from Vazyme (Vazyme). Both cell lines were maintained in Dulbecco’s minimal essential medium (DMEM) (GIBCO) supplemented with 10% fetal bovine serum (FBS) (Corning) and penicillin (100U/ml)-streptomycin (100mg/ml) (GIBCO). Clinical isolate of SARS-CoV-2 named BetaCoV/Shenzhen/SZTH-003/2020 (EPI_ISL_406594) was isolated the BALF sample of the COVID-19 patient using Vero cell in biosafety level 3 (BSL-3) laboratory^35^. The 50% tissue culture infectious dose (TCID_50_) assay was done to measure the titre of the viral stock as previously described^36,37^. In brief, Vero cells in 96-well plates were grown to 90% confluence and infected with 10-fold serial dilutions of the viral stock for 1 h at 37°C. Then the cells were overlaid with fresh DMEM supplemented with 2% FBS. At 4 days post infection, plates were assessed for the lowest dilution in which 50% of the wells exhibited cytopathology. The 50% tissue culture infectious dose (TCID_50_) values were calculated according to the Reed-Muench method^38^.

### Viral entry inhibition assay with SARS-CoV-2 pseudotyped virus

SARS-CoV-2 pseudotype neutralization assays were conducted to test the neutralizing activity of plasma from COVID-19 convalescents using pseudotyped HIV virus incorporated different variants of SARS-CoV-2 S proteins (Vazyme) as previously reported with some modification^39^. Serum samples were heat-inactivated at 56 °C for 30 min to remove complement activity, and then diluted in white, flat-bottom 96-well plates (Thermo Fisher) with a dilution of 1 in 10 with a total volume of 50μl. Then 200 TCID_50_ of SARS-CoV-2 pseudotyped particles in 50 μl were added to each well and incubated at 37°C for 1h. A total of 4 × 10^5^ ACE2-293T cells (Vazyme) in 100μl complete media were added per well and incubated for 48h at 37°C and 5% CO_2_. Firefly luciferase activity (luminescence) was measured using Bright-Light™ Luciferase Assay System (Vazyme) and a VARIOSKAN LUX Multi-Mode plate reader (Thermo) according to the manufacture’s protocols. Measurements were performed in triplicate and relative luciferase units (RLU) were then converted into neutralization percentages.

### Authentic SARS-CoV-2 microneutralization (MN) assays

Authentic SARS-CoV-2 microneutralization assays were performed as we described previously^23,24,40^. Briefly, plasma samples were heat-inactivated at 56 °C for 30 min to remove complement activity and then serially diluted in 2-folds with DMEM supplemented with 2% FBS from 1:10 to 1:5120. Diluted plasma samples were then mixed with 100 TCID_50_ of SARS-CoV-2 and incubated at 37°C for 1 hour. The mixture was added to Vero cells and incubated at 37°C and 5% CO_2_ for another 96 hours. Then cytopathic effect was evaluated and recorded using microscopy. The MN antibody titer was determined as the highest dilution with 50% inhibition of cytopathic effect.

### Enzyme-linked immunosorbent assay (ELISA)

RBD and nucleocapsid (N) specific binding immunoglobulin G (IgG) antibodies were measured according to the manufacture’s protocols (Sinobio). All plasma samples were heat inactivated at 56 °C for 30 min before use. The plasma samples diluted at 1:100 in 5% BSA was added to the high-binding ELISA plates (Corning) pre-coated with antigen (N protein or RBD) and incubated for 2 h at room temperature. Wells were then washed with PBS-T (PBS with 0.05% Tween-20), and secondary antibody was added and incubated for 1 h at room temperature. After another three washes with PBST, tetramethylben zidine (TMB) substrate (Sangon Biotech) was added at room temperature in the dark. After 15 minutes, the reaction was stopped with a 2M H_2_SO_4_ solution and then the absorbance was measured at 450 nm. A serum sample is considered positive when the OD is fourfold above background.

### Statistical analysis

Mann-Whitney U test was used to compare the antibody levels and MN titers between two groups. The Spearman rank correlation coefficient was used for linear correlation analysis between the antibody levels and MN titers. All statistical tests were calculated using SPSS 20.0 for Windows (IBM). P value less than 0.05 was considered statistically significant. The kinetics of anti-RBD IgG and MN titers during follow-up were calculated by the LOESS (locally estimated scatterplot smoothing) curve fitting polynomial regression using R software.

## Results

### Baseline characteristics of the cohort

A total of 214 laboratory confirmed COVID-19 patients with 51 individuals in severe group, 134 individuals in mild group and 29 individuals in asymptomatic group were included in this study for the analysis of antibody response. This cohort was 46.73% male with an average age of 48 years (range 2-79) (Table 1), and patients in severe group were significantly older than mild and asymptomatic groups. The body mass index (BMI) in severe group was significantly higher than the asymptomatic group. The median days of interval between onset to qPCR negative for SARS-CoV-2 was 21, and longer duration of viral shedding was found in severe and mild groups, compared with asymptomatic group. The median duration of hospitalization was 32, 20 and 14 days for the severe, mild and asymptomatic groups, respectively. The complete blood count for each patient either on the date of hospital admission or at the earliest time-point showed that expression levels of D-dimer, CRP, IL-6, PCT and LDH were significantly higher in severe and mild groups, while percentage of lymphocyte (LYM), ALB and CD4 count were significantly lower in severe group. Sequential samples were collected at time-points between 1 and 480 d.a.o (the d.a.o for the asymptomatic patients were calculated based on the days post laboratory confirmation) based on the availability, and no individuals with any further exposure history after discharge or vaccination of SARS-CoV-2 were found before the last follow-up (Table 1). A total of 411 plasma samples were collected, with 219 samples collected over 180 d.a.o and 137 samples over 360 d.a.o.

**Table 1.** Baseline characteristics and specimens of COVID-19 cases in this study.

| Characteristic | COVID-19 cases |  |  |  | P values |  |  |
| --- | --- | --- | --- | --- | --- | --- | --- |
|  | Total<br>(N=214) | Severe<br>(N=51) | Mild<br>(N=134) | Asymptomatic<br>(N=29) | S vs M | S vs A | M vs A |
| **Median age (range)** | 48 (2-79) | 62 (31-76) | 43 (2-79) | 25 (7-58) | <0.001 | <0.001 | <0.001 |
| **Age subgroup (N, %)** |  |  |  |  |  |  |  |
| <15 yr | 12 (5.6) | 0 (0) | 7 (5.2) | 5 (17.2) | NS<sup>#</sup> | 0.005 | 0.040 |
| 15-39 yr | 74 (34.6) | 4 (7.8) | 52 (38.8) | 18 (62.1) | <0.001 | <0.001 | 0.024 |
| 40-59 yr | 69 (32.2) | 15 (29.4) | 48 (36.4) | 6 (20.7) | NS<sup>#</sup> | NS<sup>#</sup> | NS<sup>#</sup> |
| ≥60 yr | 59 (27.6) | 32 (62.7) | 27 (20.1) | 0 (0) | <0.001 | <0.001 | 0.005 |
| **Male (n, %)** | 100 (46.73) | 31 (60.78) | 59 (44.03) | 10 (34.48) | 0.049 | 0.036 | NS<sup>#</sup> |
| **BMI\*** | 23.14 (21.05-25.22) | 24.3 (22.18-26.59) | 23.05 (21.30-25.11) | 20.83 (17.71-23.23) | 0.015 | <0.001 | <0.001 |
| **Co-existing chronic medical conditions (n, %)** | 68 (31.3) | 27 (52.9) | 35 (26.1) | 6 (20.7) | 0.001 | 0.005 | NS<sup>#</sup> |
| **Onset to admission, median days (Median, IQR<sup>†</sup>)** | 3 (2-7) | 5 (3-7) | 3 (1-6) | - | 0.006 | - | - |
| **Onset to antiviral treatment, median days (Median, IQR)** | 3 (1-6) | 5 (3-8) | 3 (1-6) | 1 (1-2) | 0.012 | <0.001 | <0.001 |
| **Onset to qPCR negative for SARS-CoV-2 (Median, IQR)** | 18 (12-26) | 21 (18-33) | 17 (13-25) | 9 (5-15) | <0.001 | <0.001 | <0.001 |
| **Duration of hospitalization (Median, IQR)** | 21 (16-30) | 32 (23-40) | 20 (16-27.75) | 14 (12-19) | <0.001 | <0.001 | <0.001 |
| **Laboratory results, median (Median, IQR)** |  |  |  |  |  |  |  |
| NEU ($\times 10^9/L$) | 3.12 (2.32-4.08) | 3.41 (2.29-4.34) | 2.68 (1.85-3.62) | 3.44 (2.96-4.3) | 0.013 | NS<sup>#</sup> | <0.001 |
| LYM ($\times 10^9/L$) | 1.36 (0.95-1.78) | 0.97 (0.66-1.28) | 1.42 (1.13-1.68) | 2.13 (1.59-2.61) | <0.001 | <0.001 | <0.001 |
| ALB (g/dl) | 43.4 (40.25-46.15) | 39.4 (38.1-42.6) | 43.9 (41.9-46.7) | 45.3 (43.8-46.9) | <0.001 | <0.001 | 0.004 |
| D-dimer ($\mu g/ml$) | 0.39 (0.24-0.59) | 0.56 (0.38-1.14) | 0.4 (0.24-0.57) | 0.23 (0.22-0.32) | <0.001 | <0.001 | 0.005 |
| CRP (mg/l) | 9.93 (1.87-36.15) | 44.7 (20.8-76.5) | 8.6 (3.22-24.52) | 0.62 (0.25-1.94) | <0.001 | <0.001 | <0.001 |
| IL-6 (pg/ml) | 9.25 (2.61-24.47) | 29.77 (17.98-50.84) | 11.77 (4.02-17.57) | 1.96 (1.5-2.94) | <0.001 | <0.001 | <0.001 |
| PCT (ng/ml) | 0.04 (0.03-0.07) | 0.06 (0.04-0.09) | 0.04 (0.03-0.05) | 0.02 (0.02-0.03) | <0.001 | <0.001 | 0.013 |
| LDH (U/l) | 222 (178-446.25) | 425 (223.5-664.5) | 227 (182.5-443.5) | 167 (148.5-186.5) | <0.001 | <0.001 | <0.001 |
| CD4 (count/ul) | 550 (388.5-742) | 320 (191-423.3) | 581 (454-764) | 779.5 (673.25-981.3) | <0.001 | <0.001 | 0.003 |
| **Samples collected during follow up (N, %)** | 411 | 82 (19.95) | 251 (61.07) | 78 (18.98) | - | - | - |
| $\leq 60$ d.a.o<sup>§</sup> | 116 (28.2) | 37 (45.1) | 51 (20.3) | 28 (35.9) | - | - | - |
| 61~120 d.a.o | 65 (15.8) | 4 (4.9) | 40 (15.9) | 21 (26.9) | - | - | - |
| 121~180 d.a.o | 19 (4.6) | 3 (3.7) | 12 (4.8) | 4 (5.1) | - | - | - |
| 181~240 d.a.o | 35 (8.5) | 2 (2.4) | 23 (9.2) | 10 (12.8) | - | - | - |
| 241~300 d.a.o | 16 (38.9) | 0 (0) | 12 (4.8) | 4 (5.1) | - | - | - |
| 301~360 d.a.o | 26 (6.3) | 10 (12.2) | 15 (6.0) | 1 (1.3) | - | - | - |
| 361~420 d.a.o | 73 (17.8) | 14 (17.1) | 51 (20.3) | 8 (10.3) | - | - | - |
| 421~480 d.a.o | 61 (14.8) | 12 (14.6) | 47 (18.7) | 2 (2.6) | - | - | - |
| **Any exposure to SARS-CoV-2 after discharge (N, %)** | 0 (0) | 0 (0) | 0 (0) | 0 (0) | - | - | - |
| **Vaccination of SARS-CoV-2 before last follow up (N, %)** | 0 (0) | 0 (0) | 0 (0) | 0 (0) | - | - | - |

\*Body Mass Index.
§Days after illness onset.
#Not significant.
†Inter-quartile range.

### Persistent neutralizing antibody response in COVID-19 convalescents

Firstly, the IgG response against receptor binding domain (RBD) of S glycoprotein and N proteins were measured by ELISA at 1:100 serum dilution over multiple time-points, and the neutralizing antibody titers were measured using microneutralization assay. The results of samples collected over 180 d.a.o were shown in Figure 1. Samples collected between 180 and 330 d.a.o possessed 100% positive rate for anti-RBD IgG, anti-N IgG antibody and detectable (MN titers ≥ 10) neutralizing activity against live SARS-CoV-2 (Figure 1A and 1B). However, individuals with undetectable anti-RBD, anti-N IgG and neutralizing activity were found in follow up days from 331 to 480 d.a.o, and the positive rates varied from 90.9 to 100, 90.9 to 100 and 72 to 87.5, respectively (Figure 1A and 1B). Notably, these individuals were mainly from the mild and asymptomatic groups, and the rate of individuals with undetectable neutralizing activity reached 50% (Figure 1C). Meanwhile, the distribution of MN titers obtained during 180 to 480 d.a.o showed that individuals with MN titers ≥ 160 decreased rapidly while proportion of MN titers ranging from negative to 80 increased rapidly along with time, indicating the decline of neutralizing activity in COVID-19 convalescents. Combining IgG response and MN titer measurements, higher coincidence rate of 93.19% were found between anti-RBD IgG and MN tiers than that for anti-RBD IgG and MN tiers (Figure 1E, Figure S1D), and also the significant correlations between MN titers and levels of anti-RBD IgG, especially the severe and mild groups (Figure 1F and Figure S1).

**Figure 1.**
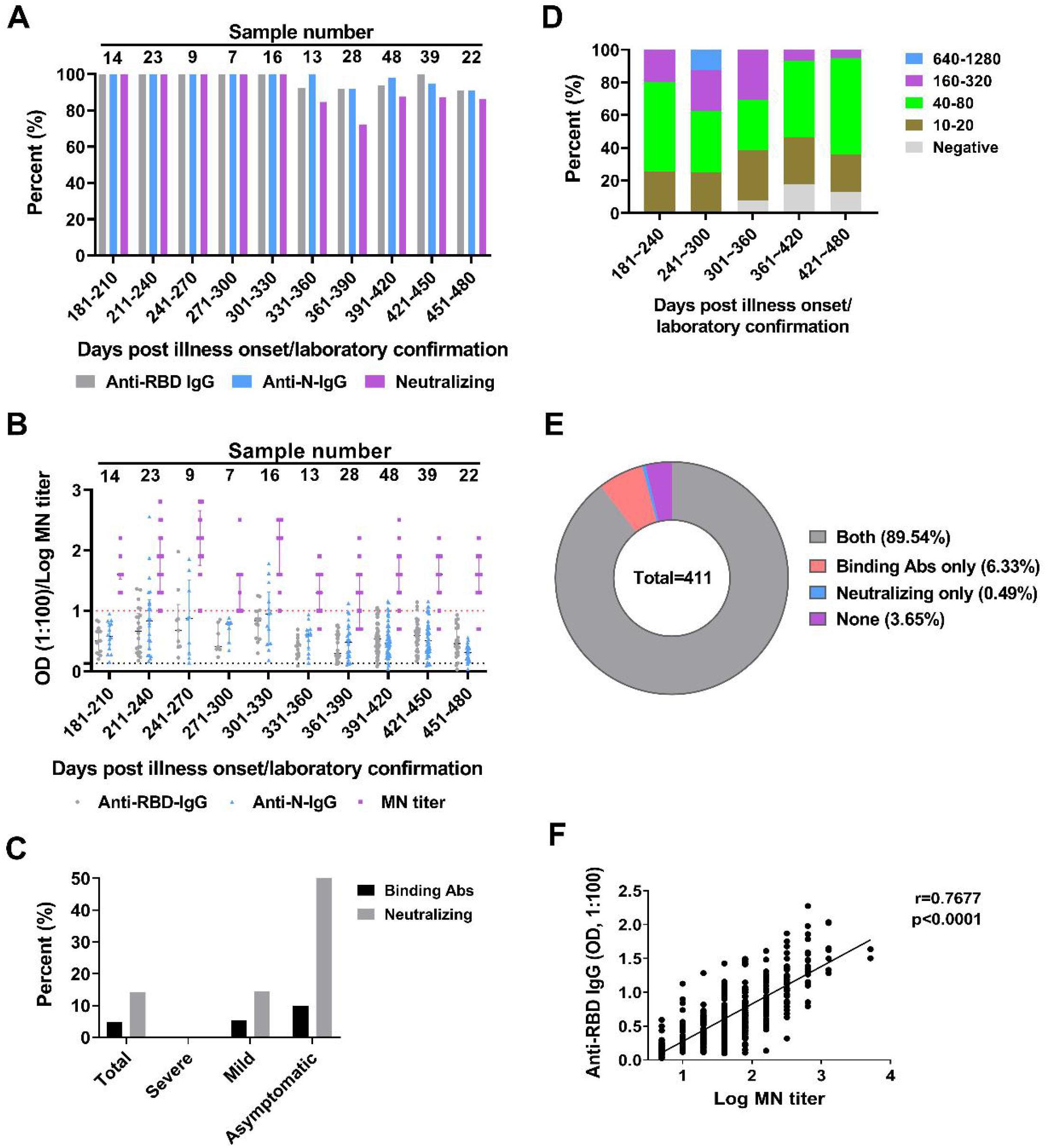
Persistent neutralizing antibody responses in COVID-19 convalescents. (A) Positive rates of the anti-RBD IgG, anti-N IgG and microneutralization (MN) titers for the 219 plasma samples from 175 COVID-19 convalescents with over 180 days follow up. (B) Detailed OD values of anti-RBD IgG, anti-N IgG and LogMN titers of the 219 plasma samples. Specimens with MN titers less than 10 were assigned a value of 5. (C) Dynamic changes of the distribution of the MN titers along with time. (D) Negative rates of anti-RBD IgG and neutralizing activity post 330 d.a.o. (E) Pie chart comparing fraction of samples with anti-RBD IgG and neutralizing activity. (F) Spearman correlation plot between the OD values of anti-RBD IgG and log MN titers. Specimens with MN titers less than 10 were assigned a value of 5.

### Comparison of neutralizing antibody response among COVID-19 patients across clinical spectrum

To determine whether disease severity impacts neutralizing antibody responses in COVID-19 patients, we firstly compared the differences of antibody development within 28 d.a.o among the severe, mild and asymptomatic groups (Figure S2). The positive rates and expression levels of both anti-RBD and anti-N IgG in severe and mild groups increased as the disease progression. For the asymptomatic patients, positive rates of both anti-RBD and anti-N IgG antibody reached 100% as early as 0∼7 days after laboratory confirmation. The positive rate of neutralizing antibody response showed similar pattern with anti-RBD and anti-N IgG antibody. Then, we further compared the available peak anti-RBD IgG levels and MN titers among the severe, mild and asymptomatic groups. Only the 48 individuals in whom the available peak anti-RBD IgG levels and MN titers occurs before the last time-point and the last time-point is ≥ 300 d.a.o were included in this analysis, with 14, 23, 11 individuals for the severe, mild and asymptomatic groups, respectively (Figure 2). Peak anti-RBD IgG levels in severe and mild groups were comparable, while significantly higher than the asymptomatic group (Figure 2A). Significant differences were found for the current anti-RBD IgG levels among the three groups, with the highest in severe group and the lowest in asymptomatic group (Figure 2C). Peak MN titers in the severe and mild groups showed no statistical significance, while both significantly higher than the asymptomatic group. In detail, peak MN titers for the severe group were all ≥ 80, and 50% belonged to range of 640-5120. For the mild group, peak MN titers varied from 10 to 1280, and 26.06% belonged to range of 640-5120. While for the asymptomatic group, all peak MN titers were ≤ 160, and 81,82% belonged to range of 10-80 (Figure 2C and 2E). As to the current MN titers, the severe group showed the highest, and no statistical significance were found between the mild and the asymptomatic groups (Figure 2D and 2F). The current MN titers of severe group mainly belonged to the range of 40-320, while negative to 40 for the mild and asymptomatic group (Figure 2F). These results indicated that COVID-19 patients with more severe disease may develop and maintain stronger neutralizing antibody response.

**Figure 2.**
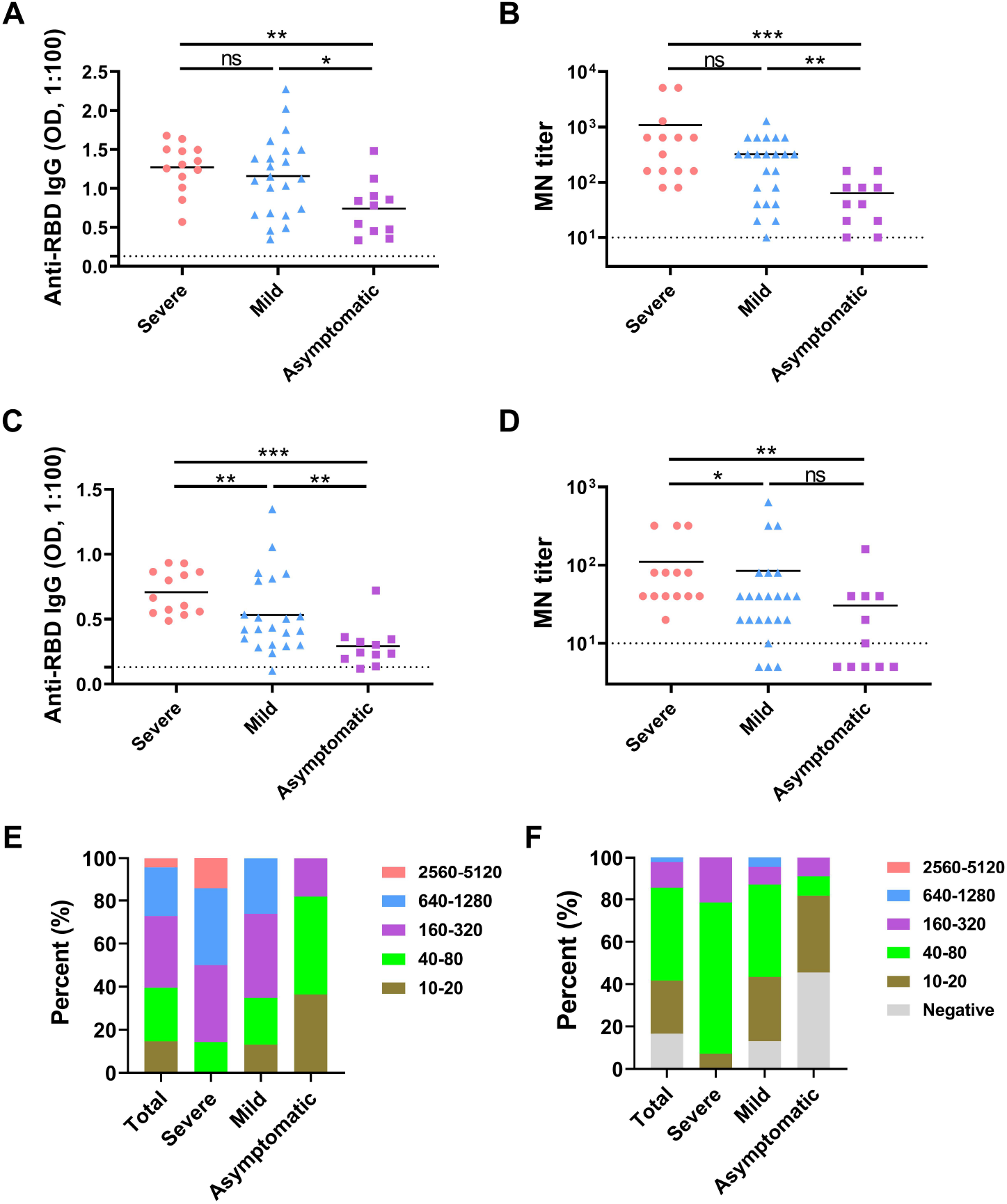
Comparison of neutralizing antibody response among different clinical spectrum of COVID-19 convalescents. (A) and (B) The available peak levels of anti-RBD IgG and MN titers for the severe, mild and asymptomatic groups. (C) and (D) The current levels of anti-RBD IgG and MN titers for the severe, mild and asymptomatic groups. Specimens with MN titers less than 10 were assigned a value of 5. (E) and (F) Distribution of the available peak and current MN titers severe, mild and asymptomatic groups.

### Dynamic change of neutralizing antibody response in COVID-19 convalescents

To obtain an overall kinetics of antibody response in COVID-19 convalescents, the anti-RBD IgG and MN titers of the 411 plasma samples from 214 were tested and analyzed as a whole, and also stratified into severe, mild and asymptomatic groups. Generally, both the anti-RBD IgG and MN titers reached the peak at around 120 days post illness onset/laboratory confirmation, and then decreased slowly and maintained relatively stable after 400 days post illness onset/laboratory confirmation (Figure 3A and 3B). When comparing among the three groups, the results showed that it took longer (about 150 days) for the severe group to reach the peak anti-RBD IgG and MN titers than the mild (about 120 days) and asymptomatic (about 80 days) groups (Figure S3). To gain a more precise dynamics of antibody response of the COVID-19 convalescents, the detailed dynamic changes of anti-RBD IgG, anti-N IgG and MN titers in patients who had measurements at more than three time points during follow up with follow up days ≥ 300 are shown in Figure 4. The profiles of both binding and neutralizing antibody dynamics varied among different individuals. For example, for Patients #01, #09, #14, #19 and #68, both the anti-RBD IgG and nAbs rose and reached the peak rapidly after infection (within 28 d.a.o), and followed by the decrease while still be detectable up to 471 d.a.o. For patients #66, #71, #72, #73, #74, #77, both the anti-RBD IgG and nAbs were detectable within 28 d.a.o and peaked at about 60-90 d.a.o, then decreased but was still be positive up to 477 d.a.o. In contrast, nAbs in patients #67, #75 from the mild group, patients #186, #193, #195 and #196 from the asymptomatic group became undetectable at 368 to 470 d.a.o, although anti-RBD IgG was still detectable in some patients (patients #68, #186, #193, #195).

**Figure 3.**
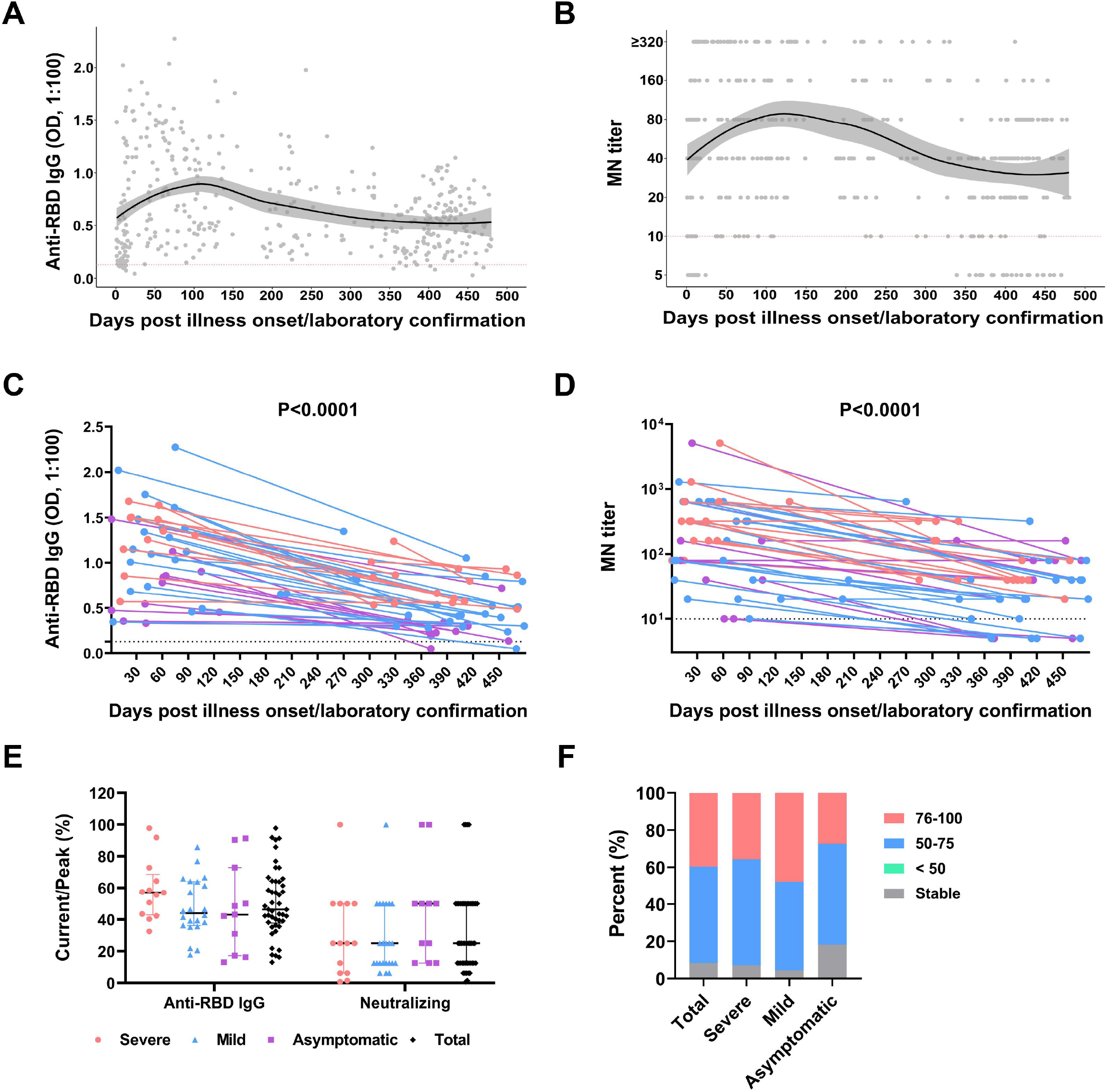
Dynamic change of neutralizing antibody response in COVID-19 convalescents. (A) and (B) Kinetics of anti-RBD IgG and MN titers against SARS-CoV-2 during the 480 days follow-up. The black solid line represents the fitted curve obtained using the LOESS (locally estimated scatterplot smoothing) curve fitting polynomial regression. Gray band areas represent 95% confidence intervals. (C) and (D) Donor-matched peak and current anti-RBD IgG/MN titers. Only individuals in whom the peak MN titers occur before the last time-point and the last time-point is > 300 d.a.o are included in this analysis (n=42), and each line represents one individual. Specimens with MN titers less than 10 were assigned a value of 5. Individuals in severe, mild and asymptomatic groups are marked in red, blue and purple, respectively. (E) The ratio of current/peak anti-RBD IgG and MN titers for the three groups. (F) Proportion of the different decrease rates in MN titers for the three groups.

**Figure 4.**
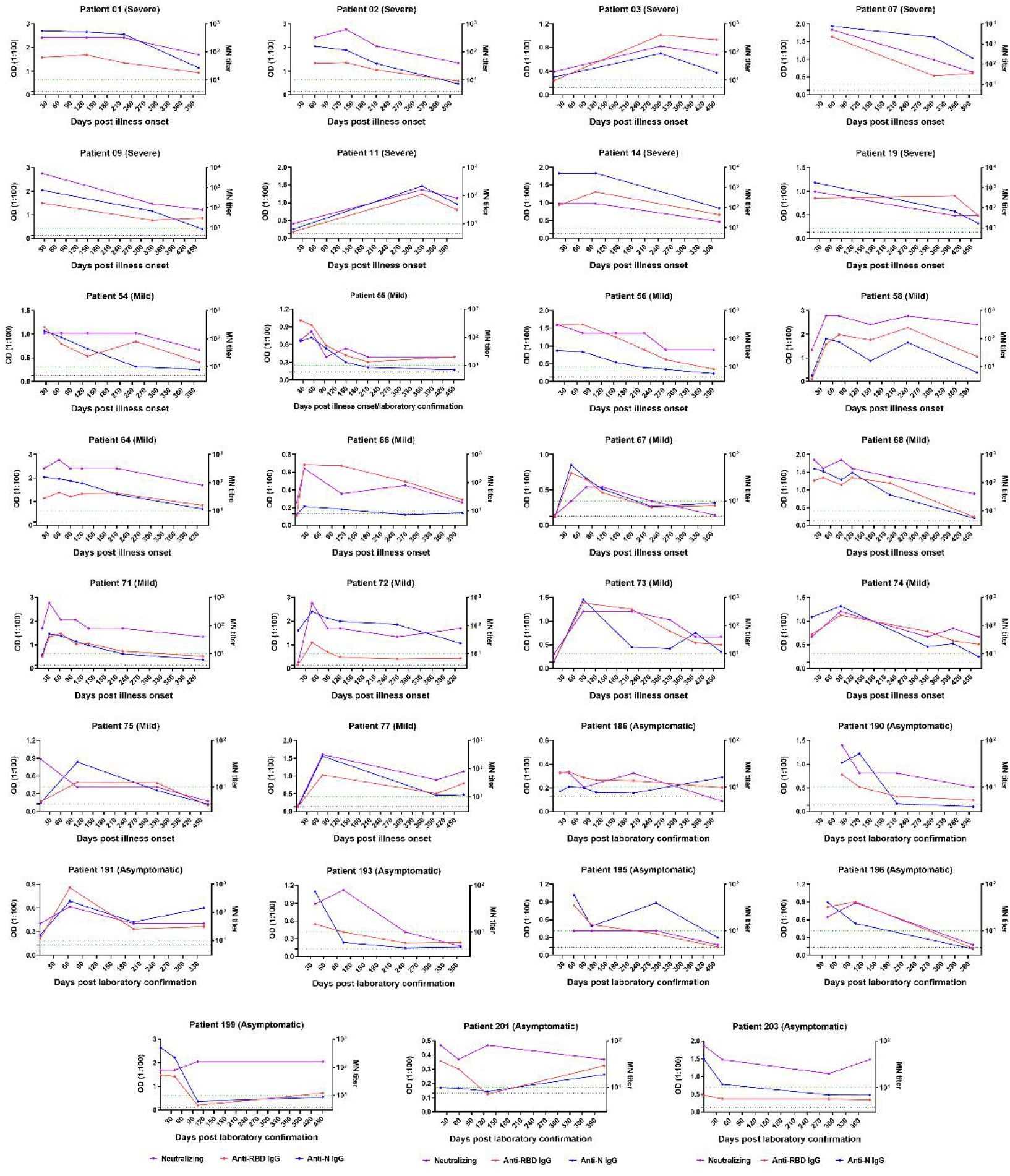
Representative dynamic changes of antibody responses (anti-RBD IgG, anti-N IgG and MN titers) in 31 COVID-19 patients during acute infection and the convalescent phase. Only patients who had measurements at more than three time points during follow up with follow up days ≥ 300 are shown. Specimens with MN titers less than 10 were assigned a value of 5.

Moreover, comparison of the peak (occurred before the last follow up) and the current (> 300 d.a.o) anti-RBD IgG levels showed that COVID-19 convalescents experienced a significant decrease in the anti-RBD IgG levels, and the median ratio of current/peak was 46.38% (range: 13.07%∼91.99%) (Figure 3C and 3E). Similar pattern was found for the MN titers (Figure 3D, 3E and 3F). A significant waning of MN titers was also found in the last follow up for 91.67% COVID-19 convalescents with ≥ 50% decrease in MN titers. Notably, stable MN titers were also found in 8.33% individuals.

### Neutralizing activity against the circulating variants of SARS-CoV-2 after one year

To test whether individuals recovered from SARS-CoV-2 infection early in the pandemic could provide protection against the circulating variants after at least one year, we tested the neutralizing activity using pseudotyped SARS-CoV-2 variants of Alpha, Beta, Gamma, Delta and Lambda (Figure 5). The information of the tested fourteen plasma samples was shown in Table S1. These samples contain samples from COVID-19 convalescents across clinical spectrum, with follow up days varied from 377 to 477 and the MN titers varied from 10 to 320. Plasma from the patients with MN titers of 160-320 (patients #47, #58, #172 and #195) could efficiently inhibit the viral entry of pseudotyped SARS-CoV-2 variants at the dilution of 1 in 10 with inhibition rates over 50%, while significantly lower inhibition rates were found in patient #172 and #195 against Beta, Delta and Lambda variants, when compared with the wild type SARS-CoV-2. Samples from patients with MN titers of 40-80 (patients #5, #20, #73, #77 and #197), 3 out of 5 could neutralize the variants of SARS-CoV-2 with inhibition rates over 50%, while very low inhibition rates against Beta, Delta and Lambda variants were found in patients #5 (MN titer 80) and #173 (MN titer 40). Meanwhile, significantly lower inhibition rates against Beta, Delta and Lambda variants were also found in all the five individuals. For the individuals with MN titers of 10-20, few individuals (only patient #13) could provide efficient protection against SARS-CoV-2 variants, while most (4/5) showed very low inhibition rates against the most variants (Alpha, Beta, Delta and Lambda), especially Beta, Delta and Lambda variants.

**Figure 5.**
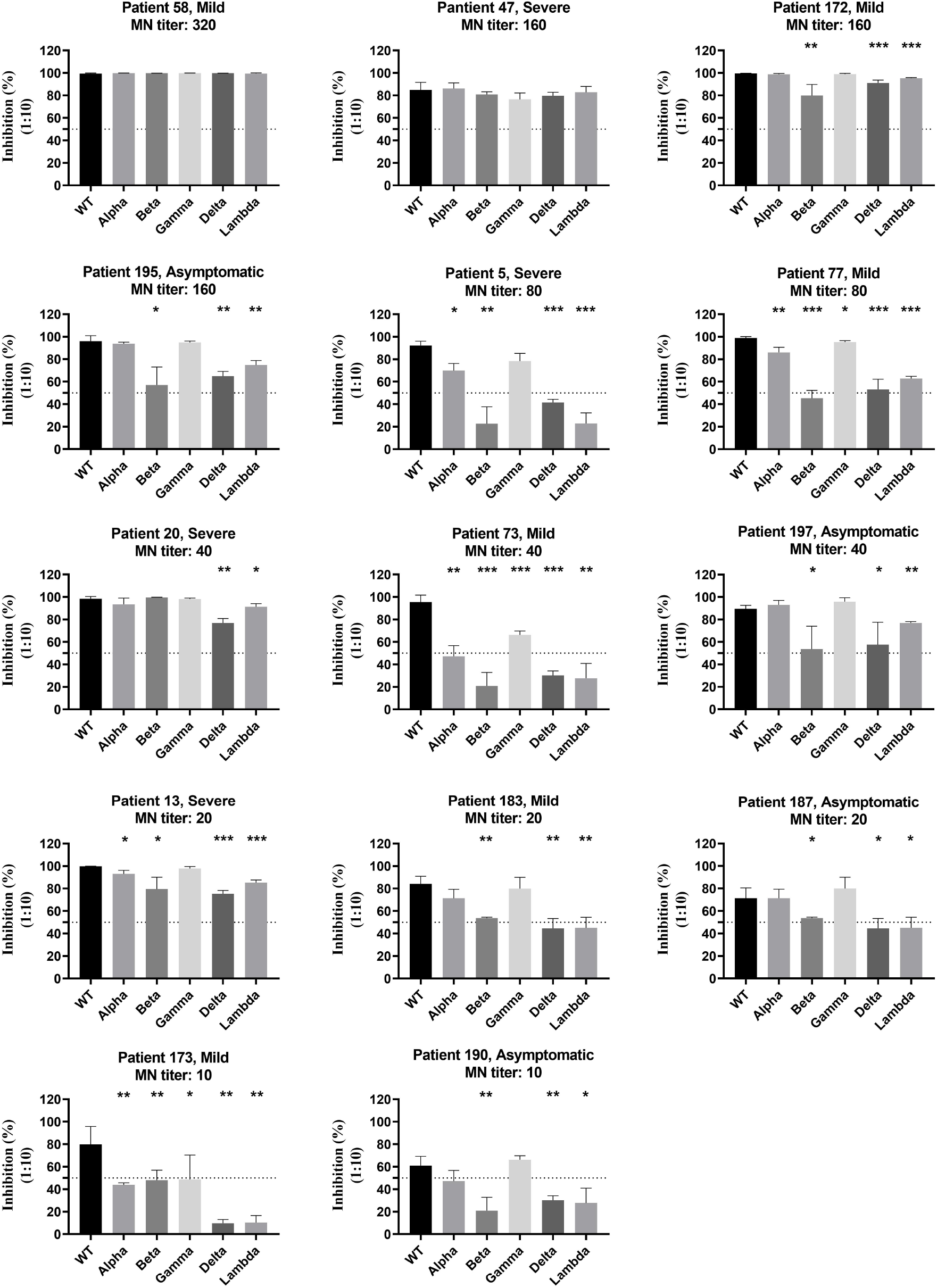
Protection of neutralizing antibody from convalescents of COVID-19 across clinical spectrum against the circulating variants of SARS-CoV-2. Neutralizing activity of 14 plasma sample collected at the last time-point (377 to 477 d.a.o) from individuals in severe, mild and asymptomatic groups were assessed using pseudotyped SARS-CoV-2 neutralization assay. The inhibition rates of single plasma against pseudotyped SARS-CoV-2 variants of Alpha, Beta, Gamma, Delta and Lambda at the dilution of 1 in 10. The belonged groups and MN titers are shown at the top of each histogram.

## Discussion

Detectable levels of neutralizing antibodies against SARS-CoV-2 have been shown to maintain positive at least 300 d.a.o for most individuals ^29^, and most studies supported that neutralizing antibody response started to decline shortly after infection, especially the mild and asymptomatic cases^14,23,25-29^. Herein, the follow up days was prolonged to 480 d.a.o, and the neutralizing activity was assessed using authentic virus based MN assay. Our results showed that neutralizing activity could be detected for all the 63 samples collected during 181∼330 d.a.o with MN titers varied from 10 to 640 (Figure 1). Individuals with undetectable neutralizing activity were found between 331 and 480 d.a.o with an overall rate of 14.29% (Figure 1C). Of concern, none of them were from the severe group, and the rate for the asymptomatic group reached 50% (Figure 1C). Distribution of MN titers obtained during 180 to 480 d.a.o showed an obvious trend of decrease along with time. Analysis of antibody dynamics showed that both the anti-RBD IgG and MN titers reached the peak at around 120 days post illness onset/laboratory confirmation, and then decreased slowly and maintained relatively stable after 400 days post illness onset/laboratory confirmation (Figure 3A and 3B) in our cohort. Moreover, when comparing the peak and current MN titers, 91.67% of the COVID-19 convalescents showed a decline over 50% in the MN titers (Figure 3F). However, it is worth noting that 8.33% individuals remained stable in MN tiers, both individuals with high (MN titer: 320) or low (MN titer: 20) MN titers (Figure 3F). These data indicated that despite of significant decline in neutralizing antibody response, it could also be maintained at least 480 days in most of the COVID-19 convalescents, while the up to 50% undetectable neutralizing activity in the asymptomatic infections is of great concern. Of note, the decline in neutralizing activity was consistent with the decline in anti-RBD IgG antibody (Figure 3E). Given the high coincidence rate (93.19%) and correlation between levels of anti-RBD IgG and the MN titers (Figure 1E and 1F), presence of the anti-RBD IgG antibody might also indicate the presence of neutralizing activity in COVID-19 convalescents for most cases.

SARS-CoV-2 reinfection and vaccine breakthrough have now been frequently reported at months after initial infection or vaccination^41-47^, suggesting that antibody mediated immunity may not confer sterilizing immunity, especially in seropositive individuals with lower amounts of baseline SARS-CoV-2 antibody^44^. Key question to address is what level of the neutralizing antibody response could provide protection against infection or reinfection. Several large cohort studies have shown that antibody-mediated immunity could provide strong protection from reinfection in seropositive individuals up to seven months after infection with protection efficacy varied from 80.5% to 95%^48-50^. A recent study suggested that the estimated neutralization level for 50% protection against detectable SARS-CoV-2 infection was 20.2% of the mean convalescent level, and the estimated neutralization level required for 50% protection from severe infection was about 3% of the mean convalescent level^51^. In such case, neutralizing antibody response was sufficient to provide enough protection against reinfection even and severe infection at 480 d.a.o (Figure 1A) for most individuals according to our study. However, SARS-CoV-2 mainly caused asymptomatic to mild infections, and asymptomatic infection constituted up to 40% of all infections^6^. Due to the high rate of undetectable neutralizing antibody response (14.41% for the mild group and 50% for the asymptomatic group) post 330 d.a.o, timely detection and vaccination might be necessary to maintain antibody mediated protection against SARS-CoV-2 for these individuals.

S protein particularly the RBD serves as the main target of neutralizing antibodies generated by SARS-CoV-2 infection and vaccination^52,53^. Studies have found that mutations in S proteins occurred frequently during the natural global transmission or under immune selection pressure and some key mutations have been found to influence the binding between S protein and its receptor or mediate the immune escape against monoclonal antibodies or convalescent plasma^54-59^. Global concerns have been raised for the potential of immune escape of antibody-dependent immunity, and fortunately recent studies have shown that the currently licensed and widely used inactivated whole virus or mRNA vaccine targeting RBD are still effective against the circulating variants, although partial resistance was observed^60-65^. In our study, all the patients were admitted to our hospital during January 11 to April 18, 2020, most likely infected by the wild type SARS-CoV-2 without the prevalent key mutations. Our results showed that antibody-dependent immunity in individuals with either high (MN titers of 160-320) or low (MN titers of 20-40) MN titers could also provide efficient protection (over 50% inhibition rates at the dilution of 1:10) against most of circulating variants during the 480 days follow up. However, significantly decreased neutralizing activities against the Beta, Delta and Lambda variants were found in most of individuals (Figure 5), which is of great concern.

Although reinfection occurred in COVID-19 convalescents or vaccinees, symptomatic reinfections and severe diseases occurred at a lower rate than primary infections^45,46,48,66^. Accordingly, it is of concern that whether COVID-19 convalescents need to receive vaccination and when, and also whether the former vaccinees need for re-vaccination and when. Currently, all the licensed vaccines have been designed to elicit immune responses to the spike protein for providing protection, therefore, it is reasonable for the assumption that the patterns of neutralizing antibody responses post-vaccination will follow the patterns seen in post-infection follow-up^67,68^. Some recent studies suggested both the binding and neutralizing antibody responses possessed similar rate of decline between vaccination of mRNA vaccine or infection during the first 7 months after vaccination^69,70^, and our study provide further hints for the durability assessment of vaccine induced antibody response. Furthermore, individuals previously diagnosed with COVID-19 could develop potent humoral immunity rapidly after a single dose of mRNA vaccine^71-73^. In our study, all the individuals of the cohort did not receive vaccination of SARS-CoV-2 before the last follow-up, while about 40 individual started to receive vaccination of inactivated SARS-CoV-2 vaccine, and the specific antibody response of these individuals merits further investigation. Moreover, giving the important role of SARS-CoV-2 specific T cell response in the control of infection, further studies based on this cohort at extended time-points of follow up are required to determine not only the longevity of the neutralizing antibody response but also the SARS-CoV-2 specific T cell response.

To our knowledge, this study is the longest follow up for the antibody dynamics of SARS-CoV-2 infection thus far, and our results provide more comprehensive insights into the natural longevity and kinetics of neutralizing antibody response in COVID-19 convalescents across clinical spectrum up to 16 months, and provide important implications for future vaccination strategies.

## Data Availability

The data underlying this article are available in the article. All data are available upon reasonable request.

## Acknowledgments

This work was supported by grants from National Natural Science Foundation of China (32170936), National Science and Technology Major Project (2021YFC0863300), the Shenzhen Science and Technology Research and Development Project (202002073000001). The authors wish to thank the biological sample bank of the Shenzhen Third People’s Hospital for bio-samples and services provided.

## Contributor

YL, JY and YY were responsible for the concept and design of the study; YY, MY, LG and YP contributed to the analysis and interpretation of data; YL, JW, LX, XL, JL, JW, ML, ZX, MZ and FW enrolled the patients and collected the clinical data; YY and MY drafted the article. YS were responsible for the critical revision for important intellectual content. All authors agree to be accountable for the content of the work.

---

**Figure S1.**
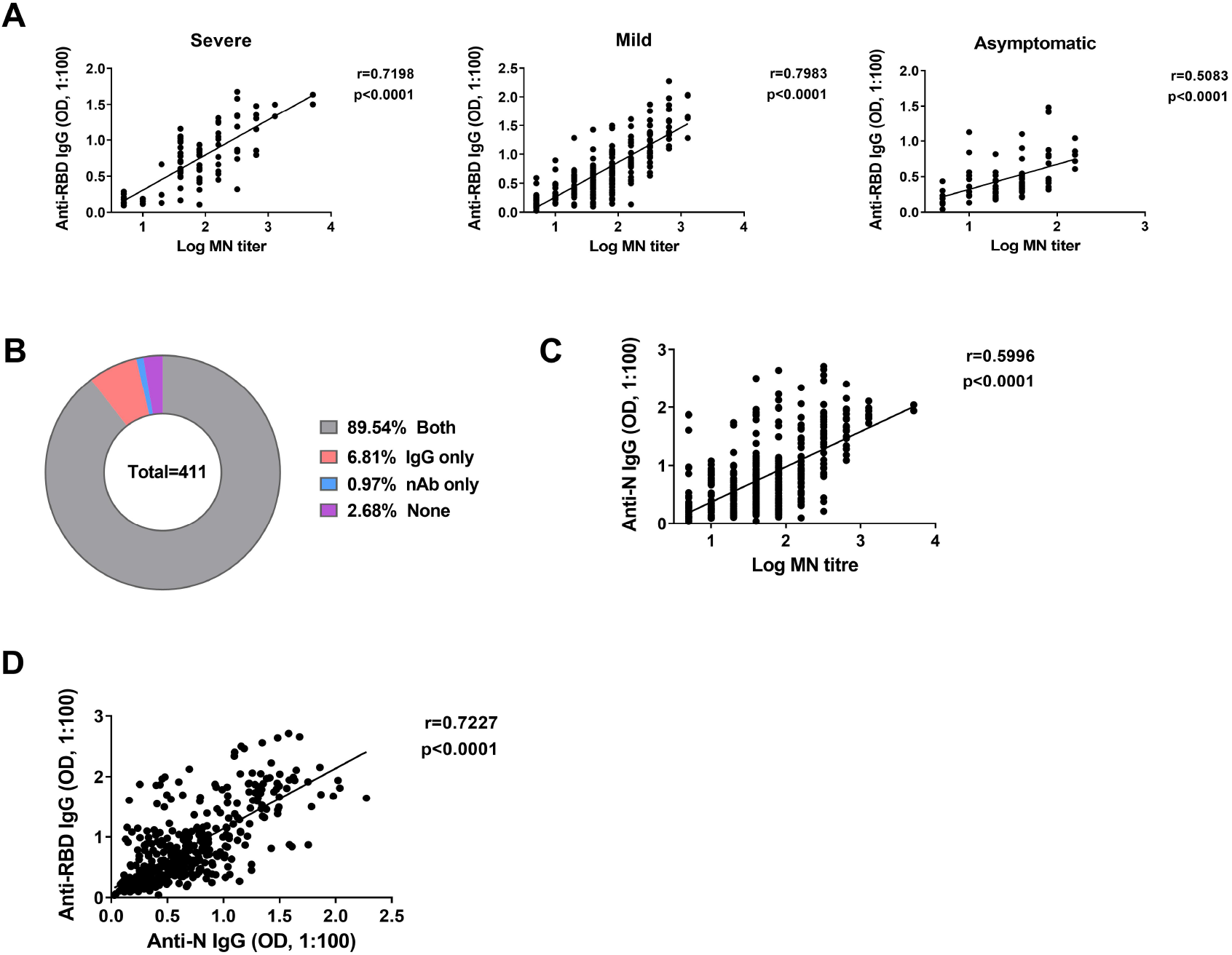
(A) Spearman correlation plot of log MN titers with anti-RBD IgG in COVID-19 patients with different clinical spectrum. Specimens with MN titers less than 10 were assigned a value of 5. (B) Pie chart comparing fraction of samples with anti-N IgG and neutralizing activity. (C) Spearman correlation plot between the OD values of anti-N IgG and log MN titers. Specimens with MN titers less than 10 were assigned a value of 5. (D) Spearman correlation plot between the OD values of anti-N IgG and anti-N IgG.

**Figure S2.**
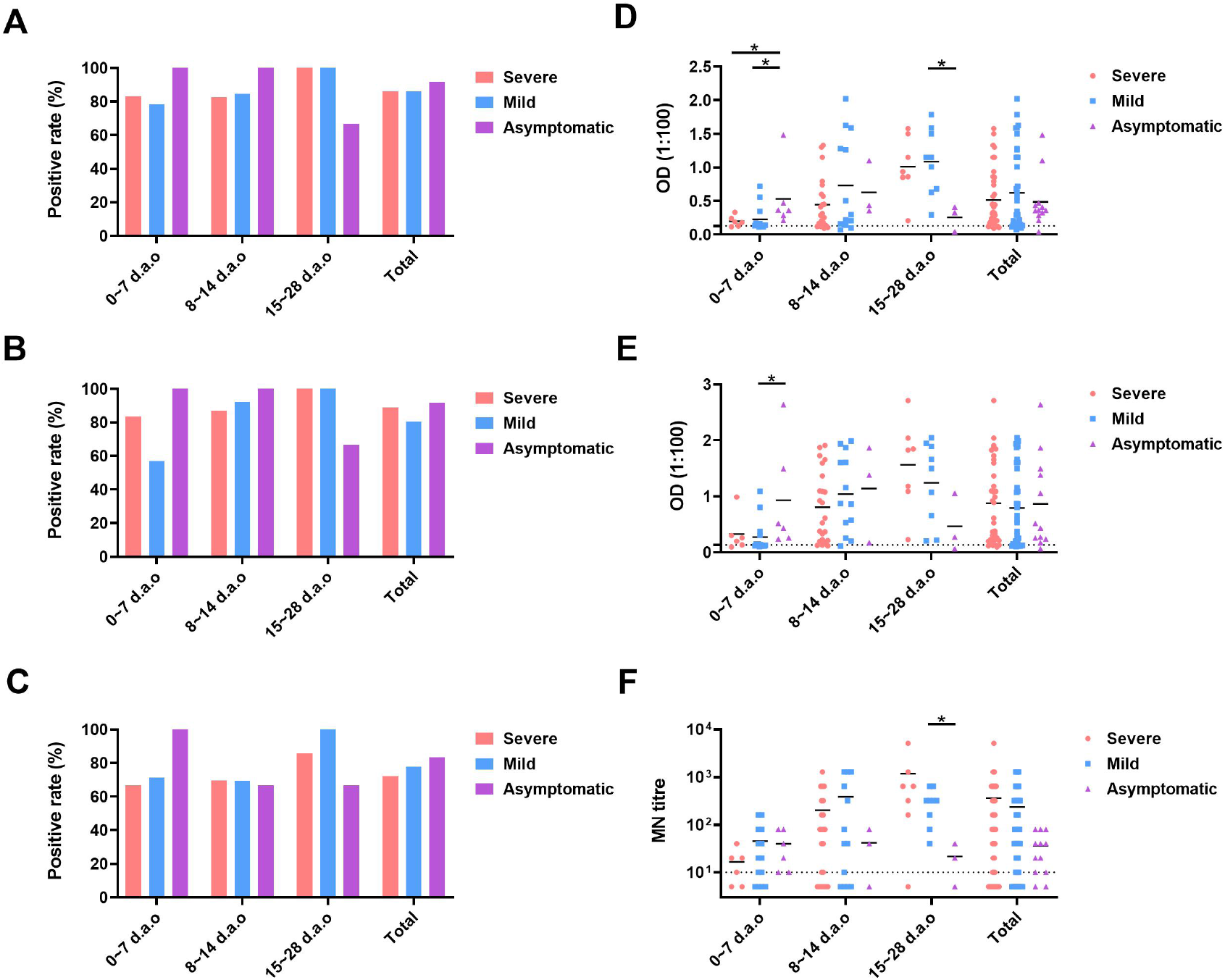
Comparison of neutralizing antibody responses of COVID-19 patients with different clinical spectrum at different stages of disease progression. (A), (B) and (C) The positive rates of anti-RBD IgG, anti-N IgG and MN titers, respectively. (D), (E) and (E) the exact values of anti-RBD IgG, anti-N IgG and MN titers, respectively.

**Figure S3.**
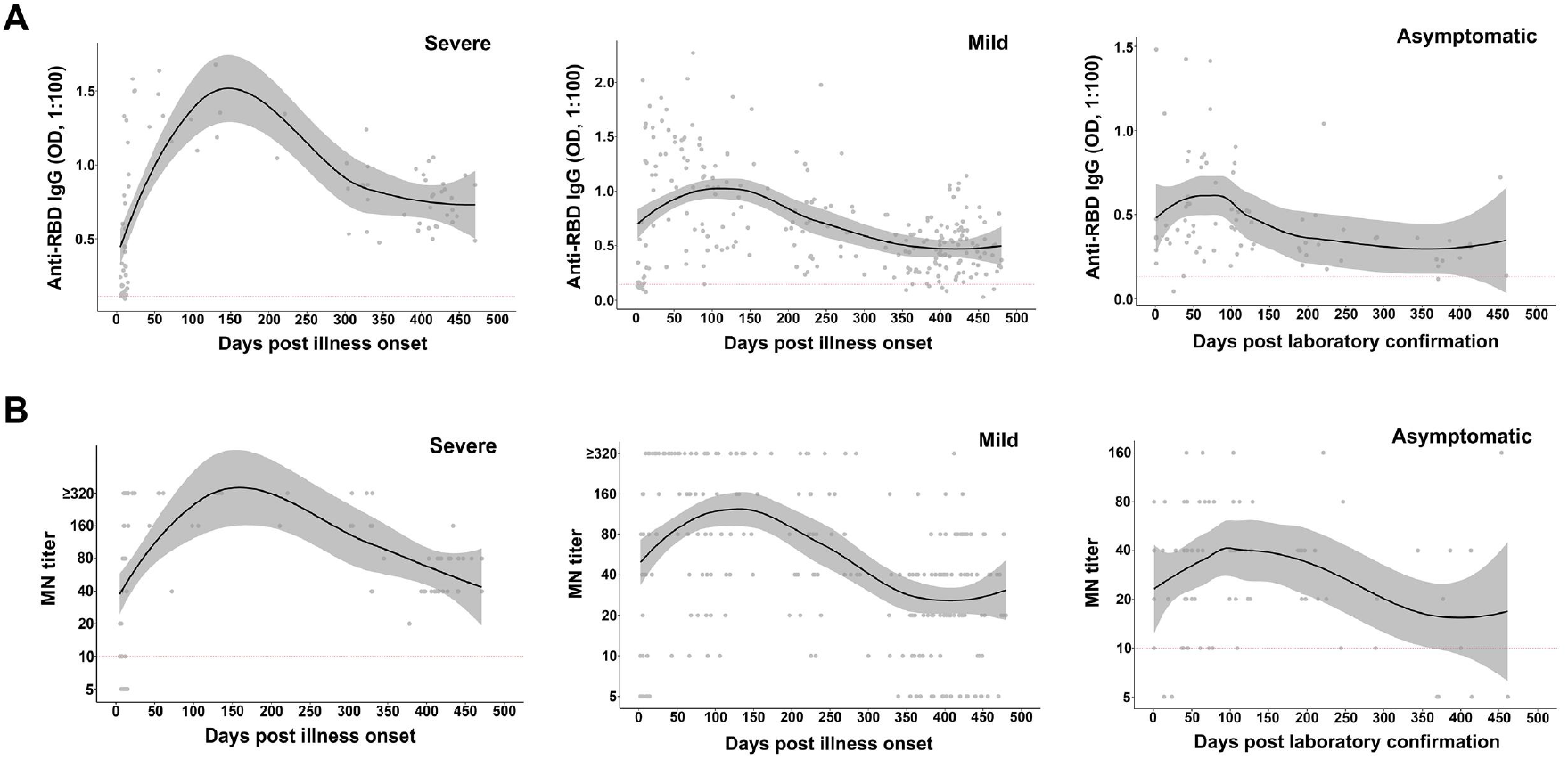
Kinetics of anti-RBD IgG (A) and MN titers (B) against SARS-CoV-2 of the COVID-19 patients with different clinical spectrum during the 480 days follow-up. The black solid line represents the fitted curve obtained using the LOESS (locally estimated scatterplot smoothing) curve fitting polynomial regression. Gray band areas represent 95% confidence intervals.

**Table S1.** Basic characteristics of plasma samples used in the pseudotyped SARS-CoV-2 neutralization assay.

| **Patient No.** | **Sex** | **Age**<br>**(range)** | **Group** | **Follow-up**<br>**days (d.a.o <sup>\*</sup>)** | **Anti-RBD**<br>**(OD<sup>§</sup>)** | **MN<sup>#</sup> titer** |
| --- | --- | --- | --- | --- | --- | --- |
| 5 | Male | 66-70 | Severe | 458 | 0.931 | 80 |
| 13 | Male | 66-70 | Severe | 378 | 0.664 | 20 |
| 20 | Female | 51-55 | Severe | 471 | 0.487 | 40 |
| 47 | Male | 46-50 | Severe | 434 | 0.662 | 160 |
| 58 | Female | 56-60 | Mild | 412 | 1.056 | 320 |
| 73 | Female | 51-55 | Mild | 470 | 0.503 | 40 |
| 77 | Female | 61-65 | Mild | 477 | 0.796 | 80 |
| 172 | Male | 56-60 | Mild | 423 | 1.018 | 160 |
| 173 | Female | 26-30 | Mild | 449 | 0.322 | 10 |
| 183 | Female | 26-30 | Mild | 477 | 0.315 | 20 |
| 187 | Male | 11-15 | Asymptomatic | 377 | 0.227 | 20 |
| 190 | Female | 31-35 | Asymptomatic | 400 | 0.243 | 10 |
| 195 | Female | 56-60 | Asymptomatic | 453 | 0.72 | 160 |
| 197 | Female | 51-55 | Asymptomatic | 413 | 0.325 | 40 |

<sup>\*</sup>Days after illness onset.
<sup>§</sup>Optical density.
<sup>#</sup>Microneutralization.

## Reference

1 Zhu, N. et al. A Novel Coronavirus from Patients with Pneumonia in China, 2019. N Engl J Med 382, 727–733, doi:10.1056/NEJMoa2001017 (2020).

2 Tan, W. et al.

3 Subramanian, R., He, Q. & Pascual, M. Quantifying asymptomatic infection and transmission of COVID-19 in New York City using observed cases, serology, and testing capacity. Proc Natl Acad Sci U S A 118, doi:10.1073/pnas.2019716118 (2021).

4 Mizumoto, K., Kagaya, K., Zarebski, A. & Chowell, G. Estimating the asymptomatic proportion of coronavirus disease 2019 (COVID-19) cases on board the Diamond Princess cruise ship, Yokohama, Japan, 2020. Euro Surveill 25, doi:10.2807/1560-7917.ES.2020.25.10.2000180 (2020).

5 Nishiura, H. et al. Estimation of the asymptomatic ratio of novel coronavirus infections (COVID-19). Int J Infect Dis 94, 154–155, doi:10.1016/j.ijid.2020.03.020 (2020).

6 Oran, D. P. & Topol, E. J. Prevalence of Asymptomatic SARS-CoV-2 Infection:A Narrative Review. Ann Intern Med 173, 362–367, doi:10.7326/m20-3012 (2020).

7 Amanna, I. J., Carlson, N. E. & Slifka, M. K. Duration of humoral immunity to common viral and vaccine antigens. N Engl J Med 357, 1903–1915, doi:10.1056/NEJMoa066092 (2007).

8 Sariol, A. & Perlman, S. Lessons for COVID-19 Immunity from Other Coronavirus Infections. Immunity 53, 248–263, doi:10.1016/j.immuni.2020.07.005 (2020).

9 Edridge, A. W. D. et al. Seasonal coronavirus protective immunity is short-lasting. Nat Med 26, 1691–1693, doi:10.1038/s41591-020-1083-1 (2020).

10 Liu, L. et al. Longitudinal profiles of immunoglobulin G antibodies against severe acute respiratory syndrome coronavirus components and neutralizing activities in recovered patients. Scand J Infect Dis 43, 515–521, doi:10.3109/00365548.2011.560184 (2011).

11 Cao, Z. et al. Potent and persistent antibody responses against the receptor-binding domain of SARS-CoV spike protein in recovered patients. Virol J 7, 299, doi:10.1186/1743-422X-7-299 (2010).

12 Choe, P. G. et al. MERS-CoV Antibody Responses 1 Year after Symptom Onset, South Korea, 2015. Emerg Infect Dis 23, 1079–1084, doi:10.3201/eid2307.170310 (2017).

13 Zhao, J. et al. Recovery from the Middle East respiratory syndrome is associated with antibody and T-cell responses. Sci Immunol 2, doi:10.1126/sciimmunol.aan5393 (2017).

14 Seow, J. et al. Longitudinal observation and decline of neutralizing antibody responses in the three months following SARS-CoV-2 infection in humans. Nat Microbiol 5, 1598–1607, doi:10.1038/s41564-020-00813-8 (2020).

15 Kim, Y. I. et al. Critical role of neutralizing antibody for SARS-CoV-2 reinfection and transmission. Emerg Microbes Infect 10, 152–160, doi:10.1080/22221751.2021.1872352 (2021).

16 Bao, L. et al. Reinfection could not occur in SARS-CoV-2 infected rhesus macaques. bioRxiv, 2020.2003.2013.990226, doi:10.1101/2020.03.13.990226 (2020).

17 Deng, W. et al. Primary exposure to SARS-CoV-2 protects against reinfection in rhesus macaques. Science 369, 818–823, doi:10.1126/science.abc5343 (2020).

18 Weinreich, D. M. et al. REGN-COV2, a Neutralizing Antibody Cocktail, in Outpatients with Covid-19. N Engl J Med 384, 238–251, doi:10.1056/NEJMoa2035002 (2021).

19 Chen, P. et al. SARS-CoV-2 Neutralizing Antibody LY-CoV555 in Outpatients with Covid-19. N Engl J Med 384, 229–237, doi:10.1056/NEJMoa2029849 (2021).

20 Duan, K. et al. Effectiveness of convalescent plasma therapy in severe COVID-19 patients. Proc Natl Acad Sci U S A 117, 9490–9496, doi:10.1073/pnas.2004168117 (2020).

21 Shen, C. et al. Treatment of 5 Critically Ill Patients With COVID-19 With Convalescent Plasma. JAMA 323, 1582–1589, doi:10.1001/jama.2020.4783 (2020).

22 Libster, R. et al. Early High-Titer Plasma Therapy to Prevent Severe Covid-19 in Older Adults. N Engl J Med 384, 610–618, doi:10.1056/NEJMoa2033700 (2021).

23 Wajnberg, A. et al. Robust neutralizing antibodies to SARS-CoV-2 infection persist for months. Science 370, 1227–1230, doi:10.1126/science.abd7728 (2020).

24 Wu, F. et al. Evaluating the Association of Clinical Characteristics With Neutralizing Antibody Levels in Patients Who Have Recovered From Mild COVID-19 in Shanghai, China. JAMA Intern Med 180, 1356–1362, doi:10.1001/jamainternmed.2020.4616 (2020).

25 Zhang, X. et al. Viral and Antibody Kinetics of COVID-19 Patients with Different Disease Severities in Acute and Convalescent Phases: A 6-Month Follow-Up Study. Virol Sin 35, 820–829, doi:10.1007/s12250-020-00329-9 (2020).

26 Bal, A. et al. Six-month antibody response to SARS-CoV-2 in healthcare workers assessed by virus neutralization and commercial assays. Clin Microbiol Infect, doi:10.1016/j.cmi.2021.01.003 (2021).

27 Weisberg, S. P. et al. Distinct antibody responses to SARS-CoV-2 in children and adults across the COVID-19 clinical spectrum. Nat Immunol 22, 25–31, doi:10.1038/s41590-020-00826-9 (2021).

28 Crawford, K. H. D. et al. Dynamics of Neutralizing Antibody Titers in the Months After Severe Acute Respiratory Syndrome Coronavirus 2 Infection. J Infect Dis 223, 197–205, doi:10.1093/infdis/jiaa618 (2021).

29 Lau, E. H. Y. et al. Neutralizing antibody titres in SARS-CoV-2 infections. Nat Commun 12, 63, doi:10.1038/s41467-020-20247-4 (2021).

30 Vanshylla, K. et al. Kinetics and correlates of the neutralizing antibody response to SARS-CoV-2 infection in humans. Cell Host Microbe 29, 917-929.e914, doi:10.1016/j.chom.2021.04.015 (2021).

31 Wang, H. et al. Dynamics of the SARS-CoV-2 antibody response up to 10 months after infection. Cell Mol Immunol 18, 1832–1834, doi:10.1038/s41423-021-00708-6 (2021).

32 Yang, Y. et al. Plasma IP-10 and MCP-3 levels are highly associated with disease severity and predict the progression of COVID-19. J Allergy Clin Immunol 146, 119–127 e114, doi:10.1016/j.jaci.2020.04.027 (2020).

33 Liu, Y. X. et al. Elevated plasma levels of selective cytokines in COVID-19 patients reflect viral load and lung injury. National Science Review 7, 1003–1011, doi:10.1093/nsr/nwaa037 (2020).

34 Long, Q. X. et al. Clinical and immunological assessment of asymptomatic SARS-CoV-2 infections. Nat Med 26, 1200–1204, doi:10.1038/s41591-020-0965-6 (2020).

35 Liu, C. et al. The Architecture of Inactivated SARS-CoV-2 with Postfusion Spikes Revealed by Cryo-EM and Cryo-ET. Structure 28, 1218–1224 e1214, doi:10.1016/j.str.2020.10.001 (2020).

36 Yang, Y. et al. Development of a quadruple qRT-PCR assay for simultaneous identification of highly and low pathogenic H7N9 avian influenza viruses and characterization against oseltamivir resistance. BMC Infect Dis 18, 406, doi:10.1186/s12879-018-3302-7 (2018).

37 Yang, Y. et al. Development of a reverse transcription quantitative polymerase chain reaction-based assay for broad coverage detection of African and Asian Zika virus lineages. Virol Sin 32, 199–206, doi:10.1007/s12250-017-3958-y (2017).

38 Reed, L. & Muench, H. A simple method for estimating fifty percent endpoints. Am. J. Hyg 37 (1938).

39 Reynolds, C. J. et al. Discordant neutralizing antibody and T cell responses in asymptomatic and mild SARS-CoV-2 infection. Sci Immunol 5, doi:10.1126/sciimmunol.abf3698 (2020).

40 To, K. K. et al. Temporal profiles of viral load in posterior oropharyngeal saliva samples and serum antibody responses during infection by SARS-CoV-2: an observational cohort study. Lancet Infect Dis 20, 565–574, doi:10.1016/S1473-3099(20)30196-1 (2020).

41 Gupta, V. et al. Asymptomatic reinfection in two healthcare workers from India with genetically distinct SARS-CoV-2. Clin Infect Dis, doi:10.1093/cid/ciaa1451 (2020).

42 To, K. K. et al. COVID-19 re-infection by a phylogenetically distinct SARS-coronavirus-2 strain confirmed by whole genome sequencing. Clin Infect Dis, doi:10.1093/cid/ciaa1275 (2020).

43 Van Elslande, J. et al. Symptomatic SARS-CoV-2 reinfection by a phylogenetically distinct strain. Clin Infect Dis, doi:10.1093/cid/ciaa1330 (2020).

44 Letizia, A. G. et al. SARS-CoV-2 seropositivity and subsequent infection risk in healthy young adults: a prospective cohort study. Lancet Respir Med 9, 712–720, doi:10.1016/s2213-2600(21)00158-2 (2021).

45 Butt, A. A. et al. Outcomes Among Patients with Breakthrough SARS-CoV-2 Infection After Vaccination: Breakthrough SARS-CoV-2 infection. Int J Infect Dis, doi:10.1016/j.ijid.2021.08.008 (2021).

46 Hacisuleyman, E. et al. Vaccine Breakthrough Infections with SARS-CoV-2 Variants. N Engl J Med 384, 2212–2218, doi:10.1056/NEJMoa2105000 (2021).

47 Kustin, T. et al. Evidence for increased breakthrough rates of SARS-CoV-2 variants of concern in BNT162b2-mRNA-vaccinated individuals. Nat Med 27, 1379–1384, doi:10.1038/s41591-021-01413-7 (2021).

48 Hall, V. J. et al. SARS-CoV-2 infection rates of antibody-positive compared with antibody-negative health-care workers in England: a large, multicentre, prospective cohort study (SIREN). Lancet 397, 1459–1469, doi:10.1016/s0140-6736(21)00675-9 (2021).

49 Abu-Raddad, L. J. et al. SARS-CoV-2 antibody-positivity protects against reinfection for at least seven months with 95% efficacy. EClinicalMedicine 35, 100861, doi:10.1016/j.eclinm.2021.100861 (2021).

50 Hansen, C. H., Michlmayr, D., Gubbels, S. M., Mølbak, K. & Ethelberg, S. Assessment of protection against reinfection with SARS-CoV-2 among 4 million PCR-tested individuals in Denmark in 2020: a population-level observational study. Lancet 397, 1204–1212, doi:10.1016/s0140-6736(21)00575-4 (2021).

51 Khoury, D. S. et al. Neutralizing antibody levels are highly predictive of immune protection from symptomatic SARS-CoV-2 infection. Nat Med 27, 1205–1211, doi:10.1038/s41591-021-01377-8 (2021).

52 Sette, A. & Crotty, S. Adaptive immunity to SARS-CoV-2 and COVID-19. Cell 184, 861–880, doi:10.1016/j.cell.2021.01.007 (2021).

53 Greaney, A. J. et al. Complete Mapping of Mutations to the SARS-CoV-2 Spike Receptor-Binding Domain that Escape Antibody Recognition. Cell Host Microbe 29, 44-57.e49, doi:10.1016/j.chom.2020.11.007 (2021).

54 Korber, B. et al. Tracking Changes in SARS-CoV-2 Spike: Evidence that D614G Increases Infectivity of the COVID-19 Virus. Cell 182, 812–827 e819, doi:10.1016/j.cell.2020.06.043 (2020).

55 Baum, A. et al. Antibody cocktail to SARS-CoV-2 spike protein prevents rapid mutational escape seen with individual antibodies. Science 369, 1014–1018, doi:10.1126/science.abd0831 (2020).

56 Greaney, A. J. et al. Comprehensive mapping of mutations in the SARS-CoV-2 receptor-binding domain that affect recognition by polyclonal human plasma antibodies. Cell Host Microbe, doi:10.1016/j.chom.2021.02.003 (2021).

57 Thomson, E. C. et al. Circulating SARS-CoV-2 spike N439K variants maintain fitness while evading antibody-mediated immunity. Cell (2021).

58 Wibmer, C. K. et al. SARS-CoV-2 501Y.V2 escapes neutralization by South African COVID-19 donor plasma. bioRxiv, doi:10.1101/2021.01.18.427166 (2021).

59 Tegally, H. et al. Emergence and rapid spread of a new severe acute respiratory syndrome-related coronavirus 2 (SARS-CoV-2) lineage with multiple spike mutations in South Africa. medRxiv, 2020.2012.2021.20248640, doi:10.1101/2020.12.21.20248640 (2020).

60 Supasa, P. et al. Reduced neutralization of SARS-CoV-2 B.1.1.7 variant by convalescent and vaccine sera. Cell 184, 2201-2211.e2207, doi:10.1016/j.cell.2021.02.033 (2021).

61 Zhou, D. et al. Evidence of escape of SARS-CoV-2 variant B.1.351 from natural and vaccine-induced sera. Cell, doi:10.1016/j.cell.2021.02.037 (2021).

62 Wang, P. et al. Antibody resistance of SARS-CoV-2 variants B.1.351 and B.1.1.7. Nature, doi:10.1038/s41586-021-03398-2 (2021).

63 Huang, B. et al. Neutralization of SARS-CoV-2 VOC 501Y.V2 by human antisera elicited by both inactivated BBIBP-CorV and recombinant dimeric RBD ZF2001 vaccines. bioRxiv, 2021.2002.2001.429069, doi:10.1101/2021.02.01.429069 (2021).

64 Planas, D. et al. Reduced sensitivity of SARS-CoV-2 variant Delta to antibody neutralization. Nature 596, 276–280, doi:10.1038/s41586-021-03777-9 (2021).

65 Lopez Bernal, J. et al. Effectiveness of Covid-19 Vaccines against the B.1.617.2 (Delta) Variant. N Engl J Med 385, 585–594, doi:10.1056/NEJMoa2108891 (2021).

66 Leidi, A. et al. Risk of reinfection after seroconversion to SARS-CoV-2: A population-based propensity-score matched cohort study. Clin Infect Dis, doi:10.1093/cid/ciab495 (2021).

67 Subbarao, K. The success of SARS-CoV-2 vaccines and challenges ahead. Cell Host Microbe 29, 1111–1123, doi:10.1016/j.chom.2021.06.016 (2021).

68 Kim, D. S., Rowland-Jones, S. & Gea-Mallorquí, E. Will SARS-CoV-2 Infection Elicit Long-Lasting Protective or Sterilising Immunity? Implications for Vaccine Strategies (2020). Front Immunol 11, 571481, doi:10.3389/fimmu.2020.571481 (2020).

69 Pegu, A. et al. Durability of mRNA-1273 vaccine-induced antibodies against SARS-CoV-2 variants. Science, doi:10.1126/science.abj4176 (2021).

70 Widge, A. T. et al. Durability of Responses after SARS-CoV-2 mRNA-1273 Vaccination. N Engl J Med 384, 80–82, doi:10.1056/NEJMc2032195 (2021).

71 Krammer, F. et al. Antibody Responses in Seropositive Persons after a Single Dose of SARS-CoV-2 mRNA Vaccine. N Engl J Med 384, 1372–1374, doi:10.1056/NEJMc2101667 (2021).

72 Demonbreun, A. R. et al. Comparison of IgG and neutralizing antibody responses after one or two doses of COVID-19 mRNA vaccine in previously infected and uninfected individuals. EClinicalMedicine 38, 101018, doi:10.1016/j.eclinm.2021.101018 (2021).

73 Manisty, C. et al. Antibody response to first BNT162b2 dose in previously SARS-CoV-2-infected individuals. Lancet 397, 1057–1058, doi:10.1016/s0140-6736(21)00501-8 (2021).

